# ADAPTIVE TIME LOCATION SAMPLING FOR COMPASS, A SARS-COV-2 PREVALENCE STUDY IN FIFTEEN DIVERSE COMMUNITIES IN THE UNITED STATES

**DOI:** 10.1101/2023.01.10.23284400

**Authors:** Sahar Z Zangeneh, Timothy Skalland, Krista Yuhas, Lynda Emel, Jean De Dieu Tapsoba, Domonique Reed, Christopher I. Amos, Deborah Donnell, Ayana Moore, Jessica Justman, the CoVPN 5002 Study Team

## Abstract

The COVPN 5002 (COMPASS) study aimed to estimate the prevalence of SARS-CoV-2 (active SARS-CoV-2 or prior SARS-CoV-2 infection) in children and adults attending public venues in 15 socio-demographically diverse communities in the United States. To protect against potential challenges in implementing traditional sampling strategies, time-location sampling (TLS) using complex sampling involving stratification, clustering of units, and unequal probabilities of selection was used to recruit individuals from neighborhoods in selected communities. The innovative design adapted to constraints such as closure of venues; changing infection hotspots; and uncertain policies. Recruitment of children and the elderly raised additional challenges in sample selection and implementation. To address these challenges, the TLS design adaptively updated both the sampling frame and the selection probabilities over time using information acquired from prior weeks. Although the study itself was specific to COVID-19, the strategies presented in this paper could serve as a case study that can be adapted for performing rigorous population-level inferences in similar settings and could help inform rapid and effective responses to future global public health challenges.

## INTRODUCTION

On March 11, 2020, the World Health Organization declared COVID-19 a pandemic. To mitigate the spread of the virus, many public and private sectors quickly adopted various forms of stay-at-home orders^1–4^. Despite these measures, the number of new infections and COVID-19-related deaths continued to rapidly rise. Community members who were unable to practice physical distancing were thus believed to be at great risk of both acquiring and transmitting the virus: This placed a disproportionate burden on individuals employed in essential industries who were afforded fewer opportunities to comply with stay-at-home orders; individuals who lived in high population-density areas who had higher risk of infection and lower access to health care resources including testing^5,6^; and individuals who regularly relied on in-person services such as transportation, childcare, food and retail. Furthermore, much of the early data of COVID-19 infections were captured through healthcare facilities, which often overrepresented people who were more likely to engage in care. Despite the high burden of COVID-19 experienced by individuals attending public venues, there was limited data that could reliably inform its magnitude. The COVID-19 Prevention Network (CoVPN) 5002, known as the COMPASS study, aimed to bridge this gap by collecting data to estimate the community-level seroprevalence of SARS-CoV-2 IgG and prevalence of SARS-CoV-2 acute infection amongst individuals 2 months of age and older, who attended public venues. The venues were in communities near 15 urban clinical research centers with diverse sociodemographic populations that were heavily burdened by the COVID-19 pandemic.

Traditional probability-based sampling methods rely on addresses, lists, or registries to sample and recruit participants^7,8^. The vast shutdowns at the beginning of the pandemic forced many well-established national probability surveys to pause or adapt new data collection strategies^9–12^, questioning the feasibility of implementing a new probability survey during the pandemic. Given logistical uncertainties in carrying out fieldwork, the COMPASS study team sought design strategies that would be more robust to changing guidelines. The COMPASS study used adaptive Time-Location Sampling (TLS)^13^ to recruit a representative random sample of individuals attending public venues. TLS was originally developed for hard-to-survey populations and assumes that individuals frequently congregate at specific locations and times^14^. TLS is a non-probability design that often used when the sampling frame is unknown. Its sampling units depend on both time and location, and the design approximates a probability sample as the number of time points and locations increase^13^, allowing approximate design-based inference^15^. The COMPASS study adapted TLS to target a broad and demographically diverse population with an unknown sampling frame. However, unlike traditional implementations of TLS, the specific locations and times of community congregation were not completely known in advance, and they needed to be updated as the study progressed. Owing to the urgency in launching this study and the changing nature of the pandemic, flexibility in the design of the study sampling frame was an important feature.

This paper describes the adaptive TLS sampling design used for the COMPASS study. The primary study outcome was based on objective blood samples that were collected by trained clinical professionals and phlebotomists, reducing measurement and processing errors in the assays^16^. To guard against nonresponse due to evolving stay-at-home guidelines, the study developed a novel adaptive sampling frame that adjusted to current closure policies and updated the selection probabilities based on enrollment information from previous weeks. To achieve desired representation of different demographic groups, unequal selection probabilities were used to oversample children, adults 60 years and older, and other community-specific hard-to-survey segments of the population. Post-stratification was used to adjust the weights on key demographic variables. To our knowledge, this study was one of the few COVID-19 studies that rigorously recruited children across diverse groups and communities.

## METHODS

### OVERVIEW OF STUDY DESIGN

Fifteen clinical research sites (CRSs) participated in the study on a voluntary basis. The sites were in Atlanta, GA; Aurora, CO; Baltimore, MD; Chicago, IL; Cincinnati, OH; Houston, TX; Miami, FL; New Orleans, LA; New York, NY (three CRSs); Newark, NJ; Philadelphia, PA; Pittsburgh, PA; and Ponce, Puerto Rico. Ethics approval was obtained from a central institutional review board (IRB) and additionally, from IRBs at participating cites, as required. Catchment areas for each site were defined as the zip code for each CRS plus all contiguous zip codes, and expanded to include additional surrounding zip codes until the required population threshold was reached^17^. The study targeted four age groups: 2 months to 17 years; 18-39 years; 40-59 years; and 60 years and older. For each site and age group, the target population was determined, as individuals attending selected community venues in their catchment area during operating hours of both the venues and the participating CRSs.

TLS was used to recruit participants from community venues. All participants went through the informed consent process. Consent for minors 17 years and younger was provided by a parent or guardian. Individuals 7 to 17 years also provided assent. Remote consent was permissible for individuals aged 15 to 17. The study collected blood serum, nasal swabs, and saliva. Dried blood spots were collected from young children and other participants who were unable to provide blood. Face-to-face interviews were used for questionnaires. The COMPASS study opened on January 11, 2021 and completed enrollments on July 31, 2021.

### CONSTRUCTION OF THE WEEKLY SAMPLING FRAME

For each catchment area, CRSs created a comprehensive roster of recruitment venues with their availabilities for each week of the study. Eligibility criteria for the venues was broad and informed by local guidelines and availability^17^. The roster was partitioned into 21 distinct and non-overlapping 4-hour time blocks per week, which we called the unrestricted Venue-Day-Time (VDT) sampling frame. The unrestricted VDT frame was then constrained to accommodate venue availability and CRS staff schedules for that week, which reduced the number of non-overlapping 4-hour time blocks for week *l* to *H*_*l*_ ≤ 21. Sites were then asked to determine the number of VDTs they could visit in each of the *H*_*l*_ 4-hour time blocks for week *l*. These numbers were informed by staff availability and the acquired information from previous weeks.

VDTs in the restricted sampling frame were flagged for (i) special events, such as farmer’s markets or food donation drives; or whether VDTs attracted (ii) children; (iii) adults over the age of 60; or (iv) hard-to-sample populations (HTS). The VDT flags could be features of venues such as playgrounds that attracted children; features of time such as food services around noon; or features of the venue and time, such as department stores offering senior discounts or religious centers where people would congregate on certain days. The VDT flags were updated each week as additional information was acquired from previous weeks.

Figure 1 describes construction of the weekly VDT frame. The weekly restricted VDT frame, the VDT flags, and the number of VDTs to be sampled in each time block, were uploaded to a web-portal managed by the central statistical and data management center two weeks in advance of sampling. Figure 2 outlines the steps taken by clinical research staff in each CRS: These steps were repeated for each site until the desired sample size was achieved for all age groups. CRSs also completed an end-of-the-day checklist that included comments on the reasons that sampled VDTs were refused (weather, policy changes, etc.) and the number of people approached in each VDT aggregated over age groups – these numbers were not disaggregated by age group as information on age would not be known prior to consent for participation.

**Figure 1.**
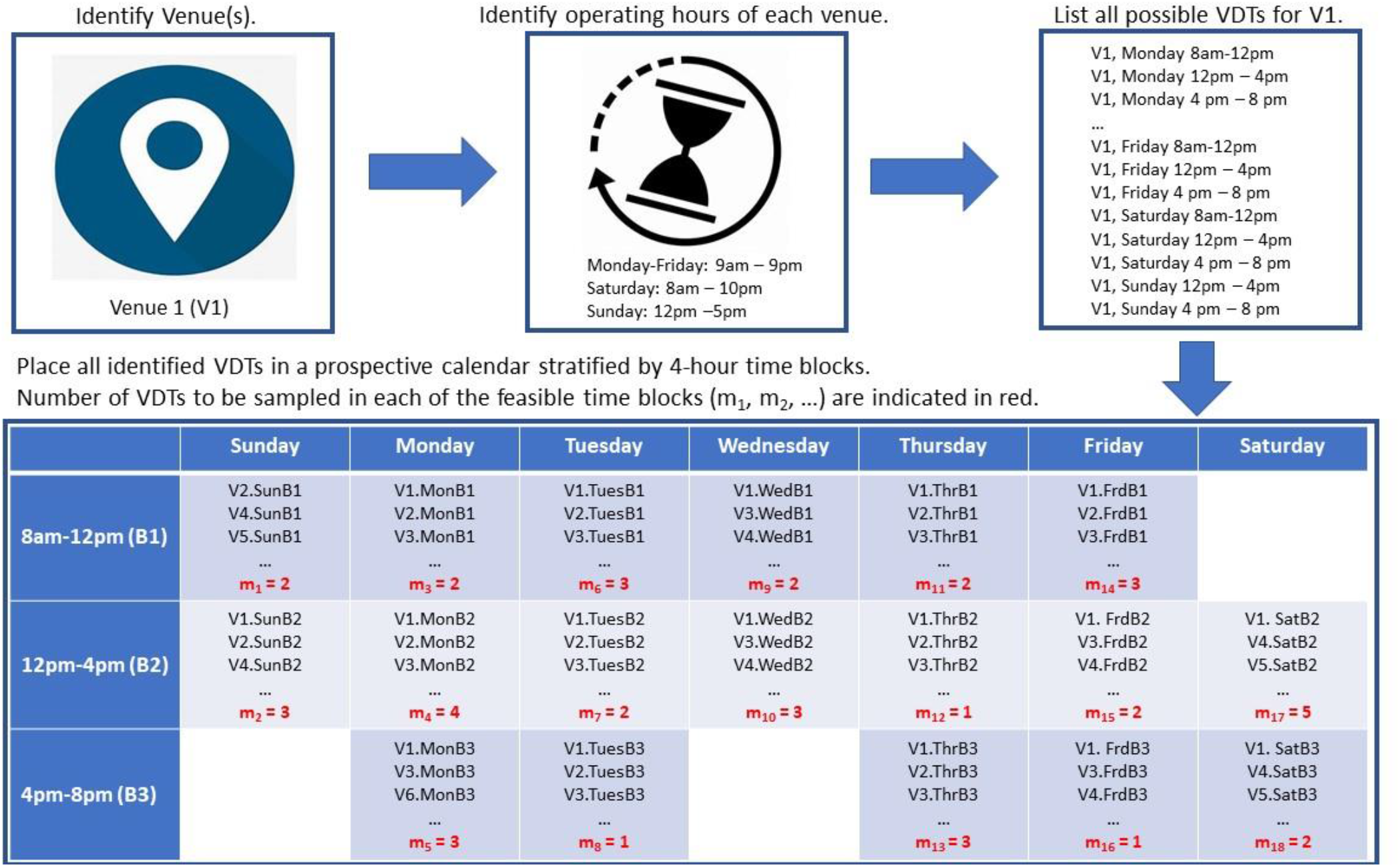
Description of the VDT sampling frame construction. Figure 1 depicts the construction of the sampling frame. The 3 square boxes on the top describe construction of the Venue-Day-Time (VDT) units corresponding to the first venue. In this example, of the maximum of the 21 strata, three are assumed to be infeasible, leaving 18 feasible time blocks. The wide rectangle in the bottom depicts the complete listings of VDTs for each of 18 strata in the restricted sampling frame for a given week. The numeric values m_1_, …, m_18_ shown in red present the number of VDTs to be sampled and visited in each of the 18 strata.

**Figure 2.**
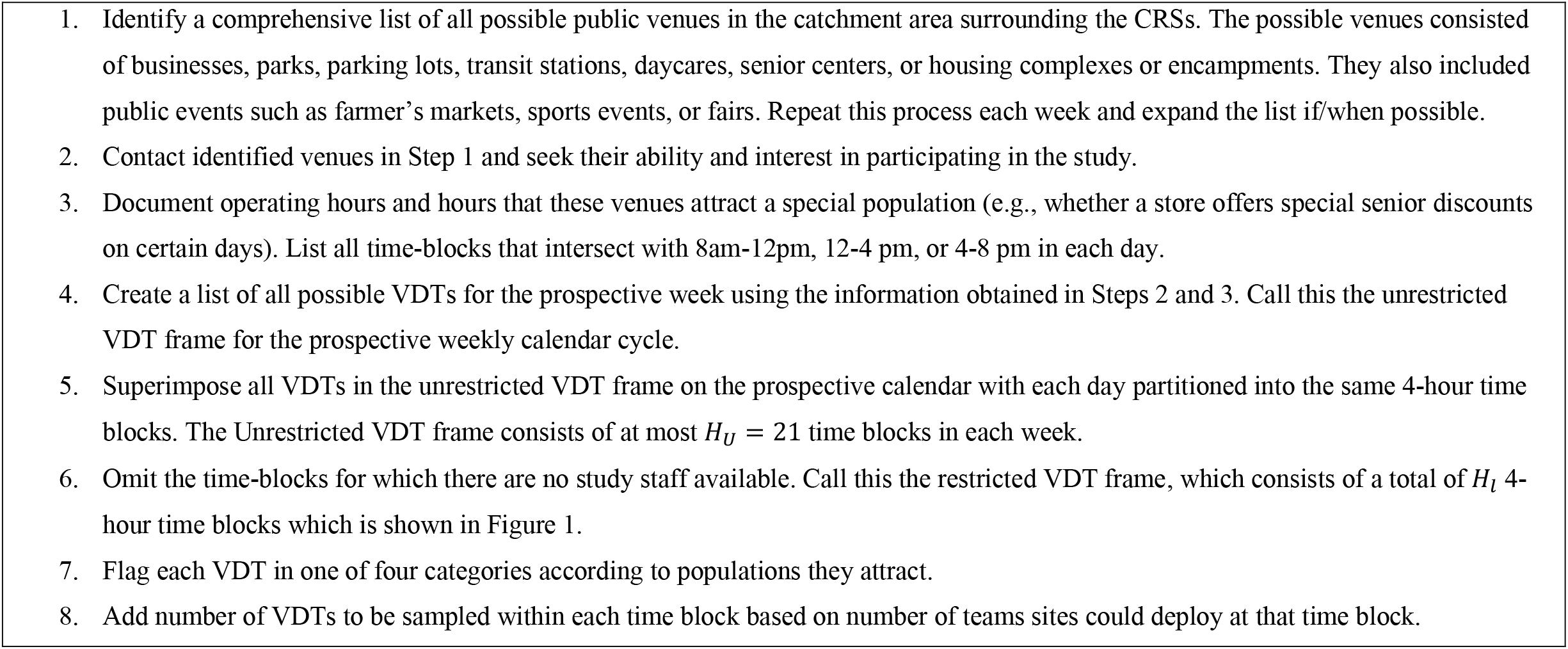
Steps for constructing the VDT sampling frame.

### TLS SAMPLING DESIGN

For each catchment area, a stratified cluster sample of VDTs was drawn, with the 4-hour time blocks in the restricted VDT frame as the sampling strata---this choice of stratification was to ease implementation. Sampling was implemented by drawing a new sample each week from that week’s restricted VDT frame and adding it to the previous sample. To avoid duplicates, prior enrollment in the study was set as an exclusion criteria^17^.

For week *l*, VDTs flagged as holding special events were sampled with a probability one. Selection probabilities for the remaining VDTs were determined using a discrete size variable 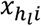 ^18–20^. The default value of 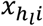 was set to 1; set to 2 for VDT(s) attracting adults over the age of 60; and set to 3 for VDTs attracting children under the age of 18. For VDTs attracting rare HTS populations, such as individuals experiencing homelessness or migrant workers, the size variable 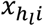 was defined as

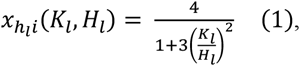

where *K*_*l*_ was the total number of strata that included an HTS in week *l*. Eq. (1) gives a size variable that is bounded between 1 and 4. This ensures that VDTs flagged as HTS get assigned the highest weights when there are only a few strata with an HTS in that week but reduces the VDT weight to 1 when many VDTs are labeled as attracting HTSs during that week. If a VDT was assigned more than one flag, its largest possible size variable was used. VDT flags were reassessed at the end of each week and used to determine the VDT flags for future weeks.

For stratum *h*_*l*_ in the restricted VDT frame for week *l*, a random sample without replacement of 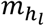 VDTs from all the 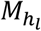 VDTs in stratum *h*_***l***_ was selected using the unequal selection probabilities described above. The value 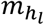 corresponded to the number of research teams deployed for data collection in that time block, which was determined based on staff availability for that week and chosen to reach prespecified weekly enrollment targets. The probability of selecting the *i*th VDT in the *h*_***l***_th stratum was thus

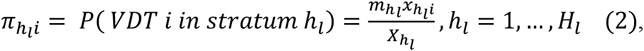

where 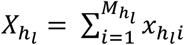 is the population total of the size variables in stratum *h*_*l*_.

At each sampled VDT, research teams approached everyone walking past the booth, handed them an informational pamphlet, and offered to answer questions, including study procedures, compensation, and return of results. Special pamphlets were designed for children. All interested participants were then recruited into the study. Random selection of individuals within VDTs was not utilized because of (i) unknown population sizes and lack of reliable projections for the expected number of individuals in the different age groups; and (ii) concerns over perceived discrimination in public settings. Because of the unknown number of individuals attending VDT *i* and the age of approached participants who chose not to enroll in the study, the selection probability of participants within each sampled VDT was estimated as

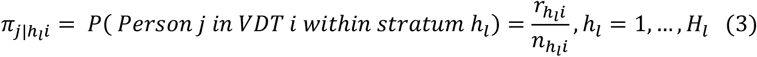

where 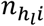 was the total number of people (regardless of age group) who were approached by CRS staff and 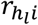 was the total number who enrolled in the study at VDT *i* in stratum *h*_*l*_, respectively. When weather became problematic or venues were unexpectedly not available because of local events or COVID restrictions, CRS staff selected from a set of randomly ordered backup VDTs to reach the desired number of VDTs. This strategy was used to mitigate missingness in VDTs and facilitate nonresponse adjustment.

### SAMPLE SIZE

The sample size was determined to achieve a prespecified margin of error (MOE) for the estimated prevalence of SARS-CoV-2 IgG seropositivity among individuals in the participating communities^21^. The MOE was chosen based on clinical relevance and was set to +/−2.5% for assumed prevalence lower than 0.05 and +/−5% for assumed prevalence greater than 0.1. A design effect of 2.5 was used to account for the clustering in the complex TLS sampling design^22–24^. Table 1 shows the sample sizes needed to achieve required MOE for a range of true infection-induced prevalence for each of the four target age groups. The sample size was first obtained for a simple random sample based on an asymptotic 95% confidence interval on the estimated prevalence and then multiplied by the design effect^25^. The target number of completed interviews for each of the four age groups was determined to be 730, which corresponded to an assumed SARS-CoV-2 IgG prevalence of 5%.

**Table 1.** Sample size requirement (per age group) to achieve +/− 5% and +/− 2.5% margin of error for a range of assumed seroprevalence after using a design effect of 2.5.

| **Assumed SARS COV-2 Prevalence** | **0.02** | **0.05** | **0.1** | **0.15** | **0.2** | **0.25** |
| --- | --- | --- | --- | --- | --- | --- |
| **+/- 5% MOE\*** | 75 | 182 | 346 | 490 | 615 | 720 |
| **+/- 2.5% MOE\*** | 301 | 730 | 1,383 | 1,959 | 2,459 | 2,881 |

\* MOE = margin of error.

### SURVEY WEIGHTING AND ANALYSIS

Combining (2) and (3) described above, the probability of selecting the *j*th individual at the *i*th VDT in stratum *h*_*l*_ was

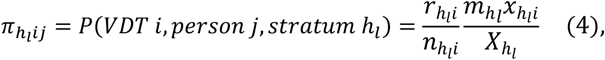

which gave a base sampling weight of 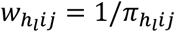. Unlike previous applications of TLS which were focused on recruiting from tightly knit populations and adjusted the weights for frequency of visits^13^, it was deemed unnecessary in this study, as the goal of the weights for frequency of visits is to achieve proportional representation, which instead achieved using post-stratification.

Raking was used to poststratify the base weights 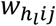 to external population margins ^26^ on race, ethnicity, and sex at birth for each of the four age categories using 2015-2019 county-level estimates from the U.S. Census Bureau’s American Community Survey ^27^. If a catchment area included two or more counties, we used average estimates from all those counties. Income and gender identity were not considered for poststratification because of their large missingness. The weights were further trimmed to bound extreme values and standardized.

## DISCUSSION

This paper presents the design of a large population-based COVID-19 seroprevalence survey in the United States. The study design facilitated robust estimation of finite population prevalence of SARS-CoV-2 acute infection and immunologic evidence of prior infection among children and adults in 15 U.S. communities. The study adapted TLS, a method often used to select for specific, hard-to-survey populations to recruit individuals from broad and diverse demographic groups in the general population. To the best of our knowledge, TLS has not previously been used to recruit individuals from the general population. One implication of this adaptation affected the construction of the survey weights, where we performed postratification instead of adjustment for frequency of visits. The study design is also novel in that it was (i) the first such survey to enroll a representative sample of community members who attended public venues; and (ii) one of the few COVID-19 surveys that directly targeted pediatric populations.

The COVID-19 pandemic created a pressing need for new, high-quality data; yet the stay-at-home policies for mitigating its spread profoundly impacted many well-established ongoing surveys such as the national health and nutrition examination survey^9–12^. Several strategies were used or proposed for sampling participants. These included using social media platforms, such as Facebook, to recruit participants^28,29^, using rapid-turnaround emails^30,31^, or proposed household surveys with mail-in testing kits^32^. Although surveys using social media or can provide large quantities of data with increased coverage of diverse populations, they suffer from significant data quality issues^33^, usually do not collect biospecimen data to capture accurate seroprevalence estimates, and are often restricted to questionnaire data or are used as only a recruitment tool^34^. Many traditional household surveys including those using email lists were also limited by their inability to collect biospecimens and suffered incredibly high nonresponse. Even proposed surveys using mail-in home testing kits were at risk of nonresponse and measurement error as the specimens were collected by study participants without supervision by study staff.

The strengths of the COMPASS study’s sampling methods include the innovative use of TLS that trades increased coverage error for reductions in nonresponse, specification, and measurement error. The sampling method was suitable for adaptation to unknown logistical and policy constraints. Limitations of the study sampling method included the nonrandom selection of the catchment areas, preventing generalizability to the national population. Another limitation is that research staff determined the venue flags, a process that may have been both subjective and operationally inefficient. Automated machine learning methods that predict the populations attending different venues by leveraging available data such as sales history, traffic patterns, or satellite images may be a promising direction for future research on alternative approaches to TLS survey methods. Finally, venues could only be used with permission from venue operators, which created some restrictions on where recruitment could take place.

Household and facility-based surveys continue to be successful for producing robust and routine estimates of the civilian non-institutionalized population. However, samples are often drawn several months ahead of field operations and can be sensitive to abrupt changes such as those experienced by COVID-19. Furthermore, such designs are limited in their ability to reach all portions of the population and often underrepresent historically marginalized populations, such as those experiencing homelessness or institutionalization. Moreover, with continuously declining response rates, probability samples are no longer shielded from various biases^35^. The TLS sampling design presented here does not replace household and facility surveys; rather, the future of surveys requires integrating multiple strategies for reaching different segments of the population. There has been growing interest in data modernization to enhance such traditional designs with nonprobability sampling designs such as TLS^36,37^. The TLS sampling design presented here can serve as an effective design for reaching historically underrepresented populations.

## CONCLUSION

Many of the challenges experienced in conducting this study were specific to the COVID-19 pandemic and the disruptions and uncertainties associated with it. However, resource limitations and unknown constraints are a constant and increasing challenge, especially in areas susceptible to natural disasters, in communities experiencing political unrest, or among individuals living in conflict zones. The methods used in this study can serve as a model that can be adapted for other studies that aim to conduct rigorous population-level assessments in such settings and thereby inform rapid and effective responses to future global public health challenges.

## Data Availability

This is a design manuscript and does not include any data analysis.

## ACKNOWLEDGEMENTS

This research was supported by the National Institute of Allergy and Infectious Diseases of the National Institutes of Health under Award UM1 AI068619. The content is solely the responsibility of the authors and does not necessarily represent the official views of the National Institutes of Health.

